# Frequency of lipoprotein(a) testing and its levels in Pakistani population

**DOI:** 10.1101/2024.02.09.24302487

**Authors:** Hijab Batool, Madeeha Khan, Quratul Ain, Omar R. Chughtai, Muhammad D. Khan, Mohammad I. Khan, Fouzia Sadiq

## Abstract

**Background:** Lipoprotein(a) [Lp(a)] is a highly atherogenic particle identified as an independent risk factor for the development of atherosclerotic cardiovascular disease (ASCVD). This study aimed to investigate the frequency of Lp(a) testing and incidence of elevated Lp(a) in the Pakistani population.

**Methods:** For this observational study, Lp(a) and lipid profile data from five years (June 2015 to October 2020) were acquired from the electronic patient records of a diagnostic laboratory (Chughtai Laboratories, Lahore). The association of age, total cholesterol (TC), high-density lipoprotein (HDL), low-density lipoprotein cholesterol (LDL-C), non-HDL, and triglyceride (TG) levels with two thresholds for Lp(a), that is, <30 mg/dL and ≥30 mg/dL, was calculated using the Kruskal Wallis test, while the association between Lp(a) levels and lipid variables was calculated using Spearman correlation.

**Results:** For five years, 1060 tests were conducted, averaging 212 tests per year. Of these tests, 37.2% showed Lp(a) levels above 30 mg/dL. There were no significant differences observed in the results between males and females. However, younger individuals displayed significantly higher Lp(a) levels. Additionally, there was only a weak correlation between Lp(a) levels and other lipid variables.

**Conclusion:** Despite being recognized as a risk factor for ASCVD in the Pakistani population, only a small proportion of the large population had their Lp(a) tested. Moreover, a significant proportion of the population lies above the threshold.

## Introduction

Lipoprotein(a) [Lp(a)] is a macromolecular structure comprising a lipid core of cholesteryl esters and triacylglycerols surrounded by an outer shell of phospholipids, free cholesterol, and apolipoprotein B-100 (apoB-100) particles linked to apolipoprotein a [apo(a)] glycoprotein^1,2^. Lp(a) is synthesized exclusively in hepatocytes and is a major carrier of oxidized phospholipids (OxPLs), which can trigger multiple pro-inflammatory pathways^3,4^. Circulating levels of Lp(a) are determined by the LPA gene locus and are not influenced by dietary or environmental factors ^5^. Data from randomized control trials suggest that diets lower in saturated fats, hormone replacement therapy (HRT) and liver disease result in lowered Lp(a) levels, whereas kidney disease results in a marked elevation of Lp(a) ^6^.

Evidence from several studies suggests that elevated Lp(a) is an independent risk factor for the development of cardiovascular diseases (CVD), including aortic valve stenosis, coronary heart disease, myocardial infarction, and stroke ^7–12^. Several international guidelines have included Lp(a) testing in their recommendations particularly for those having a high risk of cardiovascular diseases ^13–19^. The plasma Lp(a) levels are genetically determined and generally remain stable; however, genetic variability exists among different ethnic groups, with greater levels observed in Africans than in Caucasians, Hispanics, and Asian populations ^20,21^. Generally, an Lp(a) concentration of 50 mg/dL is considered a high-risk threshold ^22^. High Lp(a) levels (>50 mg/dL) were estimated to be prevalent in 20% of the population ^5^. Elevated Lp(a) has been identified as a causal risk factor in the Pakistani population; however, Lp(a) testing is not considered in routine ASCVD diagnosis ^23^.

This study aimed to investigate the incidence of Lp(a) testing and elevated Lp(a) levels in the Pakistani population.

## Methods

### Study population

For this retrospective study, anonymized Lp(a) data from the last five years (June 2015 to October 2020) was acquired from the electronic patient records of Chughtai Diagnostic Laboratories, Lahore, with collection centers across the country.

### Lp(a) and Lipid Profile Analysis

The Lp(a) levels were measured by immunoturbimetric assay (Alinity c Lp(a) kit, Product 01R1420, Abbott Laboratories, Illinois, USA). The lipid profile data for low-density lipoprotein cholesterol (LDL-C), high-density lipoprotein (HDL), total cholesterol (TC), and triglycerides (TG) of 191 subjects were measured using a homogenous assay (Abbott Alinity ci analyzer).

### Statistical analysis

The data for study variables, that is, age, Lp(a), TC, HDL, LDL, non-HDL, and TG, were reported as the minimum, maximum, median, and interquartile range (IQR). The association between Lp(a) levels and gender was calculated by using the Mann-Whitney U test. The level of significance was set at p <0.05. Differences in the significance of Lp(a) levels among different age groups were calculated using the Kruskal Wallis test. The association of age and TC, HDL, LDL, non-HDL, and TG levels with two thresholds for Lp(a), that is, <30 mg/dL and ≥30 mg/dL, was calculated using the Kruskal-Wallis test. Statistical analysis and data visualization were performed using SPSS v27.

## Results

In this study, a total of 1060 individuals were included. The details of the study’s variables, such as age, Lp(a), TC, HDL, LDL, non-HDL, and TG, are presented in Table 1. The median age of participants was 47 years. The median Lp(a) level was 20.75 (9.8-43.85) mg/dL. Among these, 37.5% (n=395) had Lp(a) levels above 30 mg/dL, while 21.3% (n=225) had Lp(a) levels above 50 mg/dL. The data for lipid variables were available for 190 individuals, where the median levels of TC, HDL, LDL, and non-HDL. have been provided (Table 1).

**Table 1:** Details of Data variables of the study.

| Data Characteristics | Minimum | Maximum | Median | IQR |
| --- | --- | --- | --- | --- |
| Age (n=1060) | 3.0 | 98.0 | 47.0 | 37.0-58.0 |
| Lp(a) (n=1060) | 1.3 | 281.3 | 20.7 | 9.8-43.8 |
| TC mg/dL (n=190) | 104.0 | 1010.0 | 192.0 | 156.7-222.5 |
| HDL-C mg/dL (n=190) | 18.0 | 86.0 | 39.0 | 32.0-46.2 |
| LDL-C mg/dL (n=190) | 50.0 | 800.0 | 123.0 | 94.7-162.2 |
| Non-HDL mg/dL (n=190) | 62.0 | 990.0 | 149.5 | 114.7-184.2 |
| TG mg/dL (n=190) | 38.0 | 891.0 | 134.5 | 97.0-215.0 |
| Lp (a) (n=190)* | 1.3 | 281.3 | 19.2 | 9.2-47.4 |
| Age (n=190)* | 3.0 | 83.0 | 41.5 | 30.7-52.0 |
| IQR- Interquartile range, Lp (a)- Lipoprotein (a), TC- Total cholesterol, TG- Triglycerides, HDL-C- High-density lipoprotein cholesterol<br>* Lp(a) levels and age data were gathered from 190 patients who had complete lipid profiles out of a larger group of 1060 patients with LP(a) data. |  |  |  |  |

The levels of Lp(a) did not show a significant difference (p=0.45) between genders. Figure 1 displays the frequency distribution of Lp(a) levels in both male and female participants. The levels of Lp(a) were compared across various age groups, including those under 20 years, between 20 to 40 years, between 40 to 60 years, and over 60 years. The results showed that there were significant differences between the groups (p=0.03) (Table 2). The median Lp(a) level for those ages below 20 years was 31.2 (9.1-92.2) mg/dL, while 23.3 (10.2-47.6) mg/dL for those aged above 60 years. Higher Lp(a) levels were observed in the younger population aged less than 20 years (Figure 2). Significant differences were observed for TC, LDL, and TG when comparing the association of study variables among the thresholds of Lp(a). There was no association found between age and HDL or non-HDL levels and Lp(a) thresholds. (Table 3).

**Table 2:**
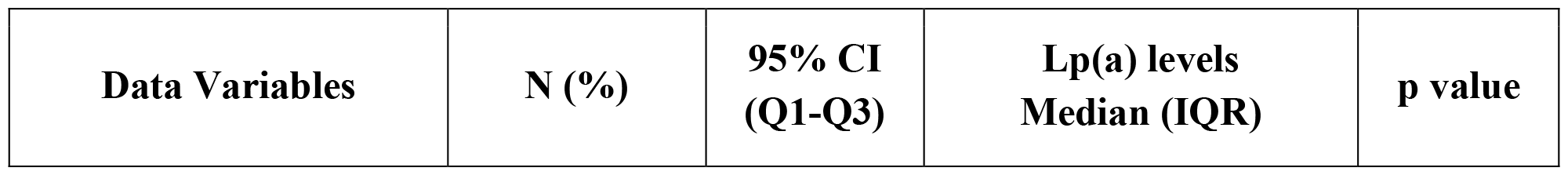
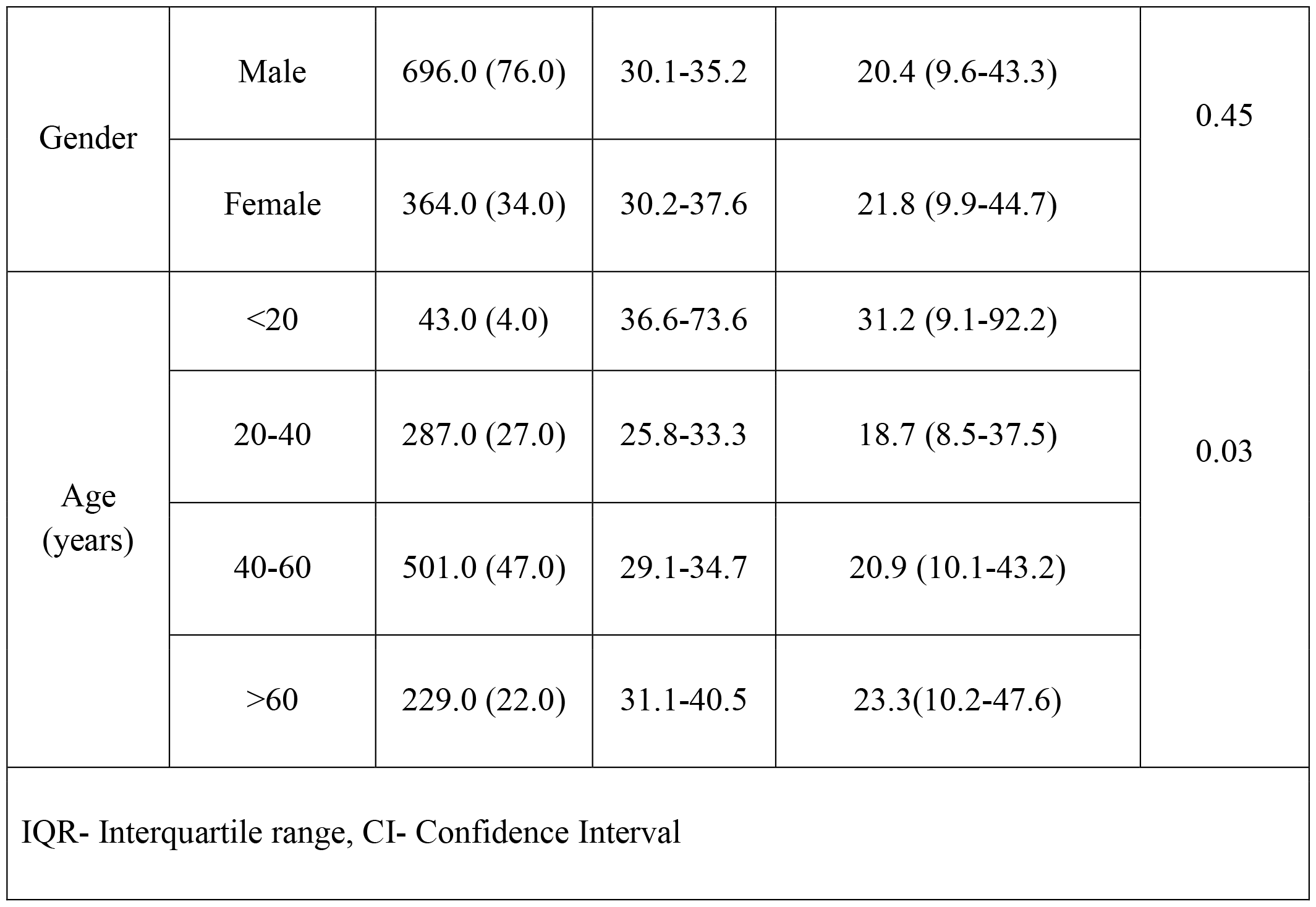
Association of gender and age with lipoprotein (a) levels.

| Data Variables | N (%) | 95% CI (Q1-Q3) | Lp(a) levels Median (IQR) | p value |
| --- | --- | --- | --- | --- |

|  |  |  |  |  |  |
| --- | --- | --- | --- | --- | --- |
| Gender | Male | 696.0 (76.0) | 30.1-35.2 | 20.4 (9.6-43.3) | 0.45 |
|  | Female | 364.0 (34.0) | 30.2-37.6 | 21.8 (9.9-44.7) |  |
| Age (years) | <20 | 43.0 (4.0) | 36.6-73.6 | 31.2 (9.1-92.2) | 0.03 |
|  | 20-40 | 287.0 (27.0) | 25.8-33.3 | 18.7 (8.5-37.5) |  |
|  | 40-60 | 501.0 (47.0) | 29.1-34.7 | 20.9 (10.1-43.2) |  |
|  | >60 | 229.0 (22.0) | 31.1-40.5 | 23.3(10.2-47.6) |  |
| IQR- Interquartile range, CI- Confidence Interval |  |  |  |  |  |

**Table 3.** Association of age, TC, HDL, LDL, non-HDL, and TG with Lp(a) thresholds i.e., <30 mg/dL and ≥30 mg/dL.

|  | **Lp(a)**<br>**<30 mg/dL (n=119)** |  | **Lp(a)**<br>**≥30 mg/dL (n=71)** |  |  |
| --- | --- | --- | --- | --- | --- |
| **N=190** | **Median** | **IQR (Q1-Q3)** | **Median** | **IQR (Q1-Q3)** | **p value** |
| Age (Years) | 42.0 | 31.0-52.0 | 40.0 | 30.0-51.0 | 0.75 |
| TC (mg/dL) | 183.0 | 150.0-216.0 | 204.0 | 172.0-239.0 | 0.01 |
| HDL-C (mg/dL) | 38.0 | 32.0-45.0 | 42.0 | 32.0-48.0 | 0.10 |
| LDL-C (mg/dL) | 114.0 | 93.0-152.0 | 135.0 | 103.0-174.0 | 0.01 |
| Non-HDL (mg/dL) | 147.0 | 112.0-181.0 | 159.0 | 124.0-194.0 | 0.07 |
| TG (mg/dL) | 142.0 | 102.0-242.0 | 115.0 | 92.0-179.0 | 0.02 |
| IQR- Interquartile range, TC- Total cholesterol, TG- Triglycerides, HDL-C- High-density lipoprotein cholesterol, LDL-C- Low density lipoprotein cholesterol |  |  |  |  |  |

**Figure 1.**
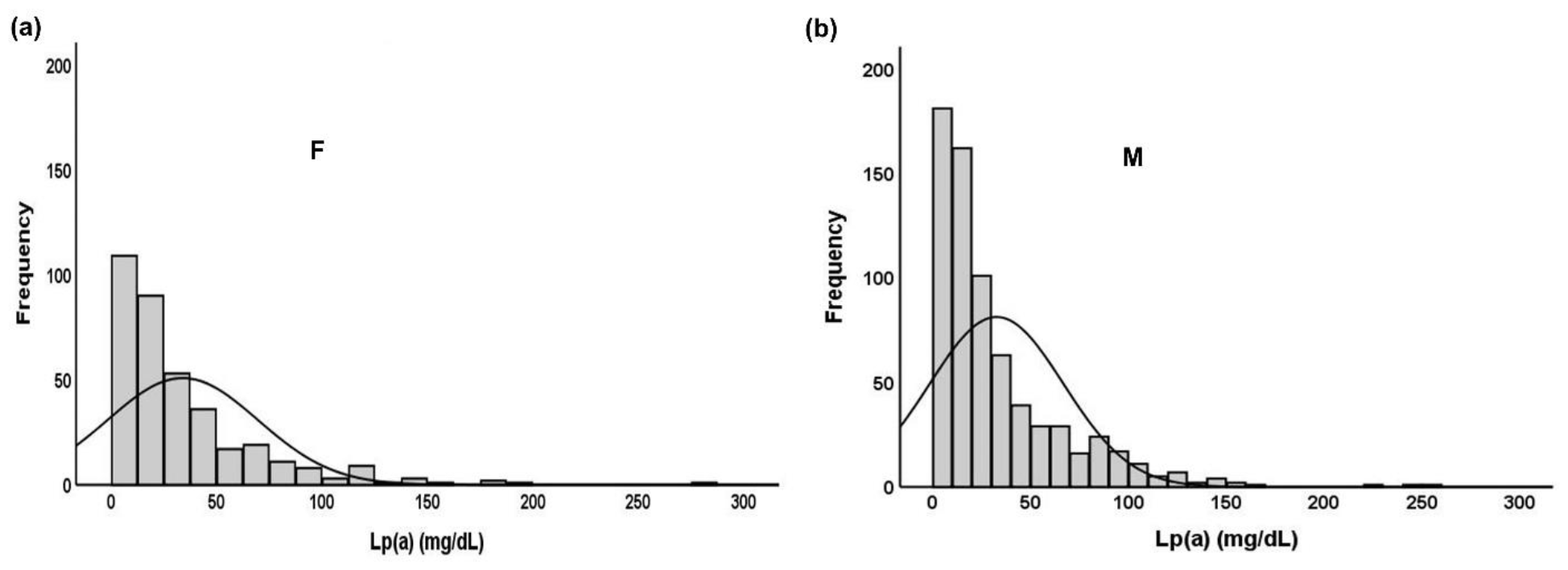
Gender-based Lp(a) frequency distribution in the Pakistani population, a)females b)males

**Figure 2.**
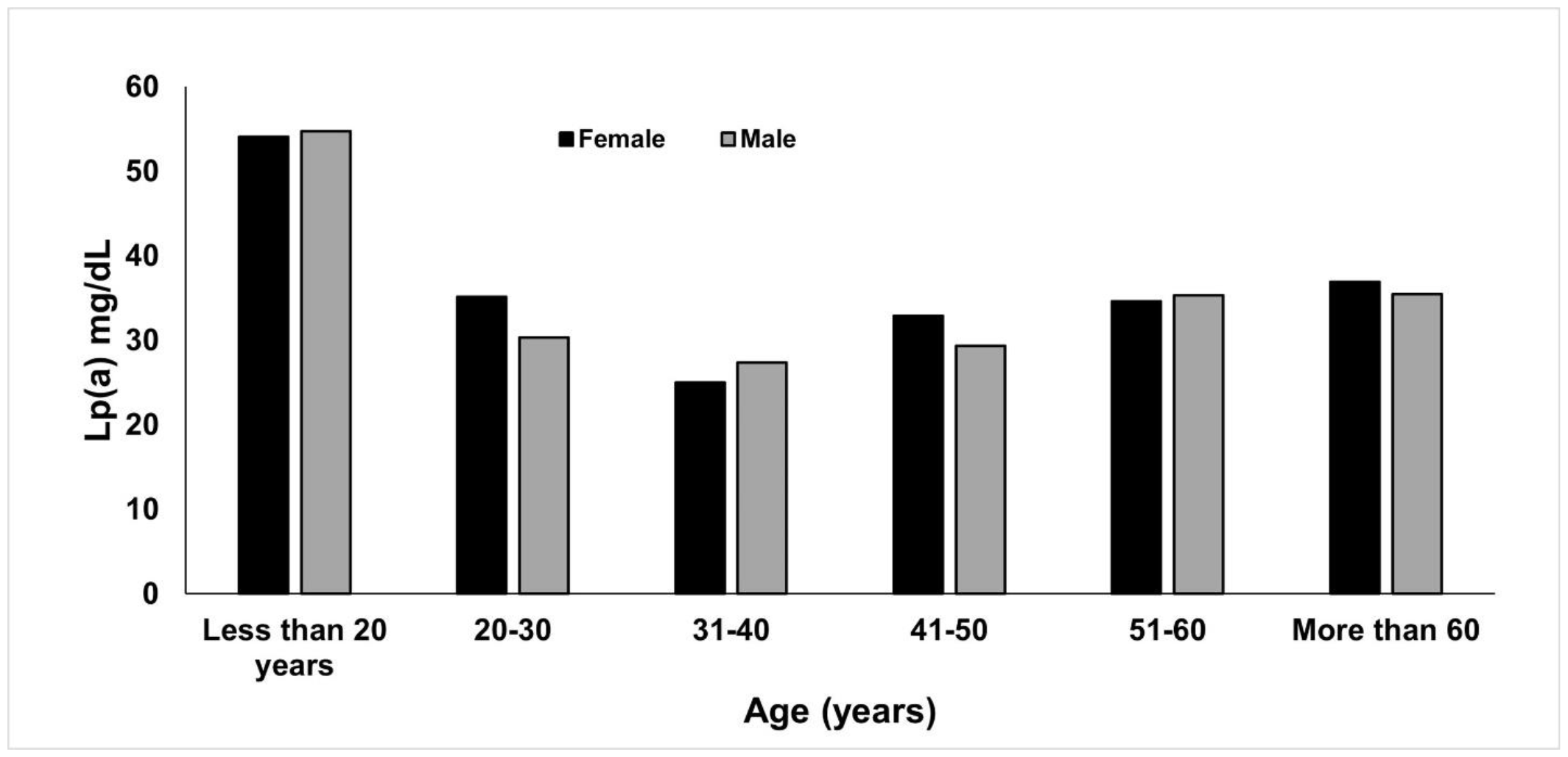
Trends of Lp(a) levels in different age groups of males and females

**Figure 3.**
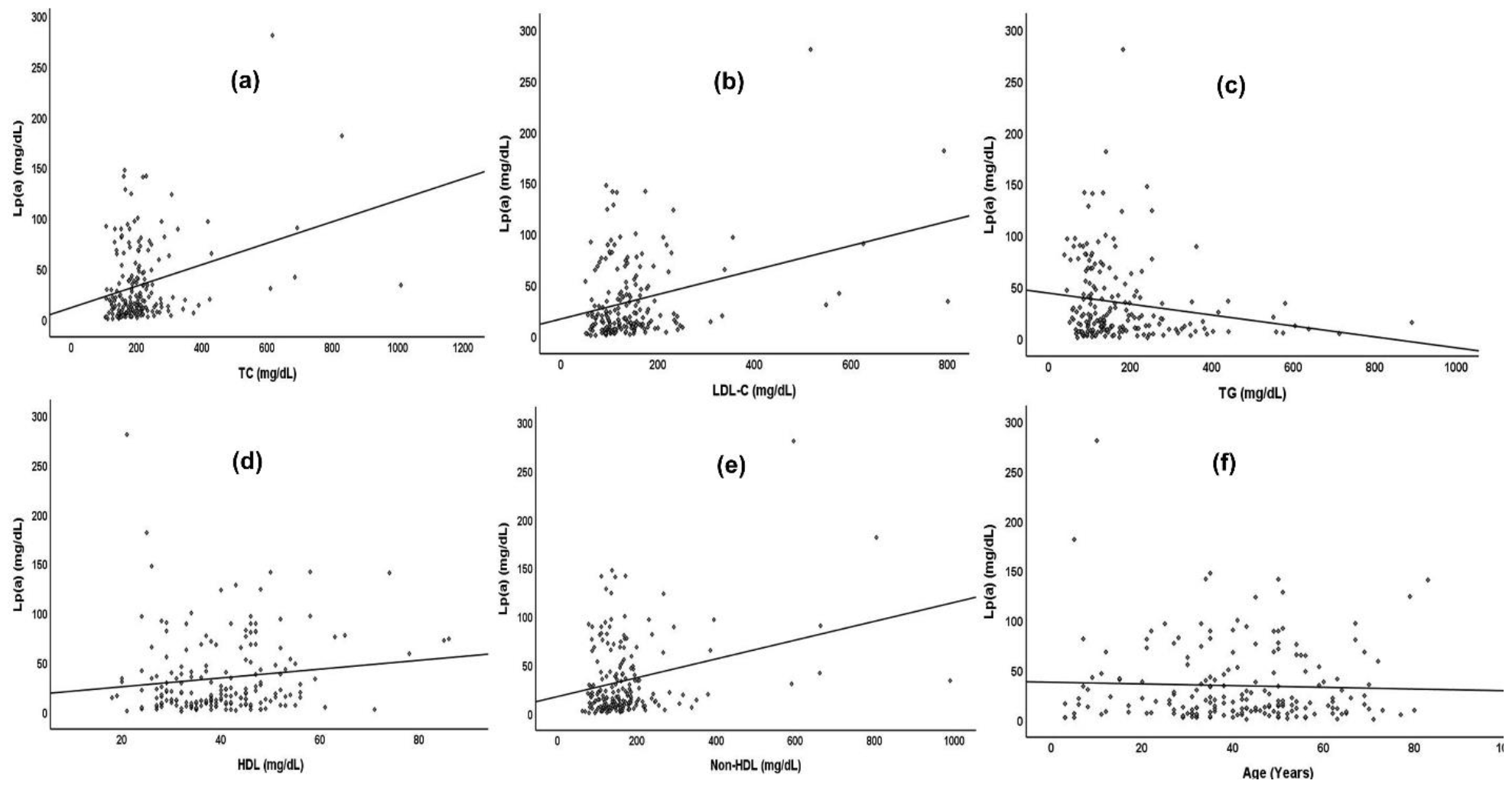
Spearman correlation between Lp(a), age, and lipid variables

## Discussion

This study presents the levels and incidence of Lp(a) testing in Pakistan. The results of the present study show that only 1060 tests were performed over five years. Among these, 21.2% had Lp(a) levels above 50 mg/dL, which is identified as a risk threshold based on the EAS/ESC guidelines, while 37.2% had Lp(a) levels above 30 mg/dL, which is categorized as a risk category according to the AHA guidelines ^14,16^.

The median Lp(a) level in the present study was 20.75 mg/dL. Previously, higher mean levels were observed for those with acute coronary syndrome (47.03 mg/dL, n=90) and diabetics (47.65 mg/dL, n=68) ^24,25^. These levels were slightly higher than those observed in the Southeast Asian population (Figure 4) ^26^. Elevated Lp(a) levels have been identified as a risk factor for coronary heart disease in the Pakistani population; however, Lp(a) testing is generally not performed routinely ^23,27^. This is also evident from the present study where only 1060 tests were booked over a span of five years in one of the largest diagnostic laboratories with centres all over Pakistan. Lp(a) testing is not highly adopted worldwide and heterogeneity regarding the incorporation of Lp(a) testing in patient care still exists ^28–31^.

**Figure 4.**
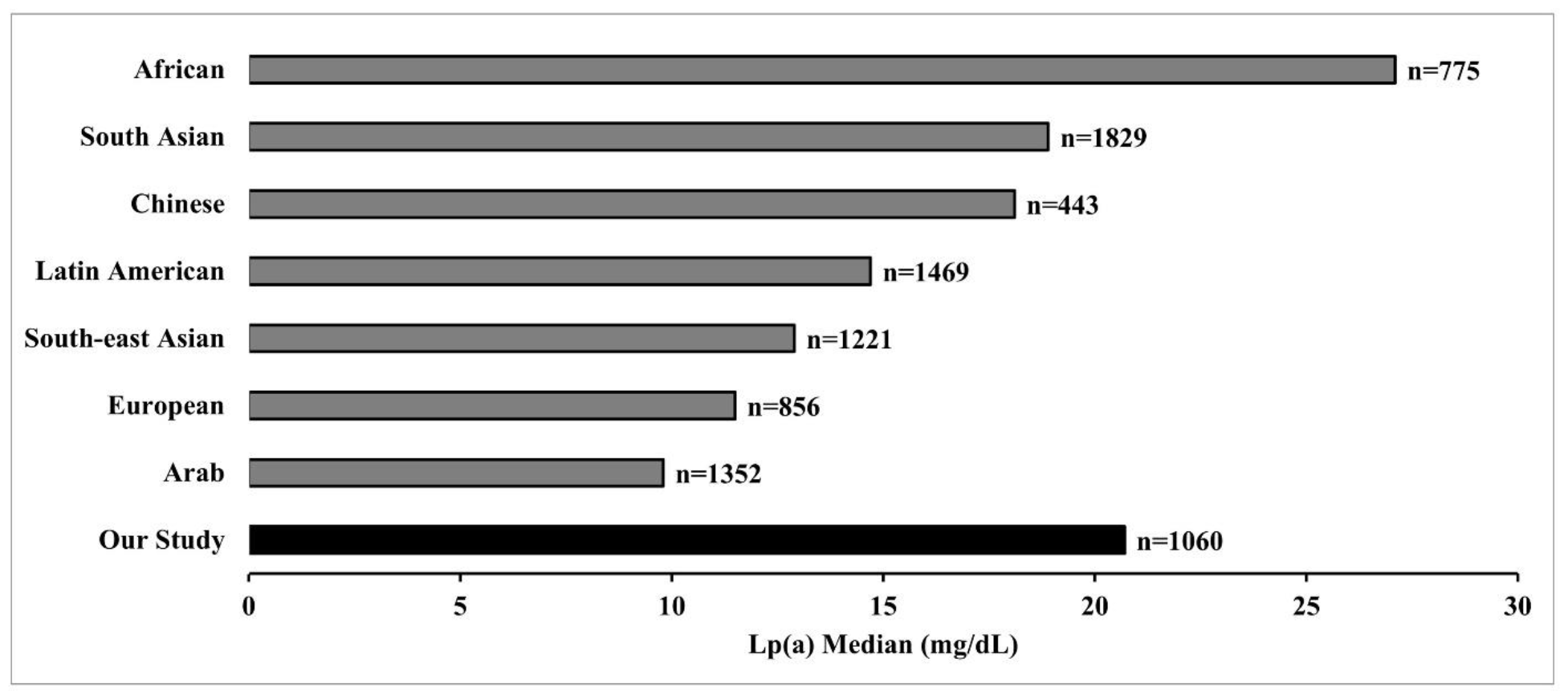
Median Lp(a) median values reported in the present study compared to the levels reported in other ethnicities

The results of the present study show that Lp(a) levels were significantly higher in younger individuals than in those aged 60 years or above. Similar results were observed in a study conducted in a multi-ethnic population with a history of ASCVD ^32^. Since elevated Lp(a) levels result in premature ASCVD, this could be the reason for higher Lp(a) observed in younger population. Moreover, the levels of Lp(a) were weakly correlated with other lipid variables such as TC, HDL, LDL-C, and TG. Previous studies have shown a weak correlation between Lp(a) levels and other lipid variables ^33,34^. However, higher Lp(a) levels were observed with higher LDL-C levels. A similar relationship was demonstrated in the studies conducted by Nicholls et al (2010) ^35^. Triglyceride levels were negatively correlated with Lp(a) levels which is consistent with the results of other studies ^34,36^. This could be either due to binding of apo(a) to the apoB of VLDL particles during the VLDL synthesis leading to the synthesis of VLDL-like particles that can be further metabolized by lipolysis or this could be due to Lp(a) metabolism by enzymes involved in TG and VLDL metabolism ^37–39^.

The major limitation of this study is that since the data were collected from a referral laboratory, details of the disease status of the individuals were not available. The details of the comorbidities and predisposition to cardiac events are unknown. Moreover, the details of the medications and their impact on lipid levels are not known.

## Conclusion

The present study showed that only a small fraction of the population is registered for Lp(a) testing. A significant proportion of our study population had Lp(a) levels above the suggested threshold. Elevated Lp(a) levels are an independent risk factor for ASCVD in the Pakistani population. Being a country with the highest rate of mortality due to cardiovascular diseases, it is imperative to screen the general population for ASCVD risk stratification, and the inclusion of Lp(a) testing can be helpful in early screening.

## Data Availability

All data produced in the present study are available upon reasonable request to the authors

## Authors’ contributions

**Conceptualization:** Fouzia Sadiq.

**Methodology:** Hijab Batool, Omar R. Chughtai, and Muhammad D. Khan.

**Resources:** Hijab Batool, Quratul Ain, Omar R. Chughtai, and Muhammad D. Khan.

**Supervision:** Mohammad I. Khan and Fouzia Sadiq.

**Validation:** Hijab Batool.

**Visualization:** Madeeha Khan and Quratul Ain.

**Writing - original draft:** Hijab Batool, Madeeha Khan, and Quratul Ain.

**Writing - review & editing:** Hijab Batool, Madeeha Khan, Quratul Ain, Omar R. Chughtai, Muhammad D. Khan, Mohammad I. Khan, and Fouzia Sadiq.

## Acknowledgements

We are thankful to Mr. Amjad Nawaz, Shifa Tameer e Millat University, Islamabad, for helping in conducting statistical analysis.

## References

1. Gaubatz JW, Heideman C, Gotto Jr AM, Morrisett JD, Dahlen GH. Human plasma lipoprotein [a]. Structural properties. Journal of Biological Chemistry. 1983;258(7):4582–4589. doi:10.1016/S0021-9258(18)32663-2

2. Jawi MM, Frohlich J, Chan SY. Lipoprotein(a) the Insurgent: A New Insight into the Structure, Function, Metabolism, Pathogenicity, and Medications Affecting Lipoprotein(a) Molecule. Jiang XC, ed. J Lipids. 2020;2020:3491764. doi:10.1155/2020/3491764

3. Schmidt K, Noureen A, Kronenberg F, Utermann G. Structure, function, and genetics of lipoprotein (a). J Lipid Res. 2016;57(8):1339–1359. doi:10.1194/jlr.R067314

4. McLean JW, Tomlinson JE, Kuang WJ, Eaton DL, Chen EY, Fless GM, Scanu AM, Lawn RM. cDNA sequence of human apolipoprotein(a) is homologous to plasminogen. Nature. 1987;330(6144):132–137. doi:10.1038/330132a0

5. Tsimikas S. A Test in Context: Lipoprotein(a): Diagnosis, Prognosis, Controversies, and Emerging Therapies. J Am Coll Cardiol. 2017;69(6):692–711. doi:10.1016/j.jacc.2016.11.042

6. Enkhmaa B, Berglund L. Non-genetic influences on lipoprotein(a) concentrations. Atherosclerosis. 2022;349:53–62. doi:10.1016/j.atherosclerosis.2022.04.006

7. Bhatia HS, Ma GS, Taleb A, Wilkinson M, Kahn AM, Cotter B, Yeang C, DeMaria AN, Patel MP, Mahmud E, Reeves RR, Tsimikas S. Trends in testing and prevalence of elevated Lp(a) among patients with aortic valve stenosis. Atherosclerosis. 2022;349:144–150. doi:10.1016/j.atherosclerosis.2022.01.022

8. Zhu L, Zheng J, Gao B, Jin X, He Y, Zhou L, Huang J. The correlation between lipoprotein(a) elevations and the risk of recurrent cardiovascular events in CAD patients with different LDL-C levels. BMC Cardiovasc Disord. 2022;22(1):1–10. doi:10.1186/S12872-022-02618-5/TABLES/4

9. Cao J, Steffen BT, Budoff M, Post WS, Thanassoulis G, Kestenbaum B, Mcconnell JP, Warnick R, Guan W, Tsai MY. Lipoprotein(a) levels are associated with subclinical calcific aortic valve disease in white and black individuals: The multi-ethnic study of atherosclerosis. Arterioscler Thromb Vasc Biol. 2016;36(5):1003–1009. doi:10.1161/ATVBAHA.115.306683

10. Clarke R, Peden JF, Hopewell JC, Kyriakou T, Goel A, Heath SC, Parish S, Barlera S, Franzosi MG, Rust S, Bennett D, Silveira A, Malarstig A, Green FR, Lathrop M, Gigante B, Leander K, de Faire U, Seedorf U, Hamsten A, Collins R, Watkins H, Farrall M. Genetic variants associated with Lp(a) lipoprotein level and coronary disease. N Engl J Med. 2009;361(26):2518–2528. doi:10.1056/NEJMOA0902604

11. Arora P, Kalra R, Callas PW, Alexander KS, Zakai NA, Wadley V, Arora G, Kissela BM, Judd SE, Cushman M. Lipoprotein(a) and Risk of Ischemic Stroke in the REGARDS Study. Arterioscler Thromb Vasc Biol. 2019;39(4):810–818. doi:10.1161/ATVBAHA.118.311857

12. Kumar P, Swarnkar P, Misra S, Nath M. Lipoprotein (a) level as a risk factor for stroke and its subtype: A systematic review and meta-analysis. Scientific Reports 2021 11:1. 2021;11(1):1–13. doi:10.1038/s41598-021-95141-0

13. Cegla J, Neely RDG, France M, Ferns G, Byrne CD, Halcox J, Datta D, Capps N, Shoulders C, Qureshi N, Rees A, Main L, Cramb R, Viljoen A, Payne J, Soran H. HEART UK consensus statement on Lipoprotein(a): A call to action. Atherosclerosis. 2019;291:62–70. doi:10.1016/j.atherosclerosis.2019.10.011

14. Grundy SM, Stone NJ, Bailey AL, Beam C, Birtcher KK, Blumenthal RS, Braun LT, de Ferranti S, Faiella-Tommasino J, Forman DE, Goldberg R, Heidenreich PA, Hlatky MA, Jones DW, Lloyd-Jones D, Lopez-Pajares N, Ndumele CE, Orringer CE, Peralta CA, Saseen JJ, Smith SC, Sperling L, Virani SS, Yeboah J. 2018 AHA/ACC/AACVPR/AAPA/ABC/ACPM/ADA/AGS/APhA/ASPC/NLA/PCNA Guideline on the Management of Blood Cholesterol: A Report of the American College of Cardiology/American Heart Association Task Force on Clinical Practice Guidelines. J Am Coll Cardiol. 2019;73(24):e285–e350. doi:10.1016/J.JACC.2018.11.003

15. Handelsman Y, Jellinger PS, Guerin CK, Bloomgarden ZT, Brinton EA, Budoff MJ, Davidson MH, Einhorn D, Fazio S, Fonseca VA, Garber AJ, Grunberger G, Krauss RM, Mechanick JI, Rosenblit PD, Smith DA, Wyne KL. Consensus Statement by the American Association of Clinical Endocrinologists and American College of Endocrinology on the Management of Dyslipidemia and Prevention of Cardiovascular Disease Algorithm – 2020 Executive Summary. Endocrine Practice. 2020;26(10):1196–1224. doi:10.4158/CS-2020-0490

16. Mach F, Baigent C, Catapano AL, Koskinas KC, Casula M, Badimon L, Chapman MJ, De Backer GG, Delgado V, Ference BA, Graham IM, Halliday A, Landmesser U, Mihaylova B, Pedersen TR, Riccardi G, Richter DJ, Sabatine MS, Taskinen MR, Tokgozoglu L, Wiklund O, Mueller C, Drexel H, Aboyans V, Corsini A, Doehner W, Farnier M, Gigante B, Kayikcioglu M, Krstacic G, Lambrinou E, Lewis BS, Masip J, Moulin P, Petersen S, Petronio AS, Piepoli MF, Pinto X, Raber L, Ray KK, Reiner Z, Riesen WF, Roffi M, Schmid JP, Shlyakhto E, Simpson IA, Stroes E, Sudano I, Tselepis AD, Viigimaa M, Vindis C, Vonbank A, Vrablik M, Vrsalovic M, Gomez JLZ, Collet JP, Windecker S, Dean V, Fitzsimons D, Gale CP, Grobbee DE, Halvorsen S, Hindricks G, Iung B, Jüni P, Katus HA, Leclercq C, Lettino M, Merkely B, Sousa-Uva M, Touyz RM, Nibouche D, Zelveian PH, Siostrzonek P, Najafov R, Van De Borne P, Pojskic B, Postadzhiyan A, Kypris L, Spinar J, Larsen ML, Eldin HS, Strandberg TE, Ferrières J, Agladze R, Laufs U, Rallidis L, Bajnok L, Gudjonsson T, Maher V, Henkin Y, Gulizia MM, Mussagaliyeva A, Bajraktari G, Kerimkulova A, Latkovskis G, Hamoui O, Slapikas R, et al. 2019 ESC/EAS Guidelines for the management of dyslipidaemias: Lipid modification to reduce cardiovascular risk. Eur Heart J. 2020;41(1):111–188. doi:10.1093/eurheartj/ehz455

17. Pearson GJ, Thanassoulis G, Anderson TJ, Barry AR, Couture P, Dayan N, Francis GA, Genest J, Grégoire J, Grover SA, Gupta M, Hegele RA, Lau D, Leiter LA, Leung AA, Lonn E, Mancini GBJ, Manjoo P, McPherson R, Ngui D, Piché ME, Poirier P, Sievenpiper J, Stone J, Ward R, Wray W. 2021 Canadian Cardiovascular Society Guidelines for the Management of Dyslipidemia for the Prevention of Cardiovascular Disease in Adults. Canadian Journal of Cardiology. 2021;37(8):1129–1150. doi:10.1016/J.CJCA.2021.03.016

18. Stefanutti C, Julius U, Watts GF, Harada-Shiba M, Cossu M, Schettler VJ, De Silvestro G, Soran H, Van Lennep JR, Pisciotta L, Klör HU, Widhalm K, Moriarty PM, D’Alessandri G, Bianciardi G, Bosco G, De Fusco G, Di Giacomo S, Morozzi C, Mesce D, Vitale M, Sovrano B, Drogari E, Ewald N, Gualdi G, Jaeger BR, Lanti A, Marson P, Martino F, Migliori G, Parasassi T, Pavan A, Perla FM, Brunelli R, Perrone G, Renga S, Ries W, Romano N, Romeo S, Pergolini M, Labbadia G, Di Iorio B, De Palo T, Abbate R, Marcucci R, Poli L, Ardissino G, Ottone P, Tison T, Favari E, Borgese L, Shafii M, Gozzer M, Pacella E, Torromeo C, Parassassi T, Berni A, Guardamagna O, Zenti MG, Guitarrini MR, Berretti D, Hohenstein B, Saheb S, Bjelakovic B, Williams H, De Luca N. Toward an international consensus—Integrating lipoprotein apheresis and new lipid-lowering drugs. J Clin Lipidol. 2017;11(4):858–871.e3. doi:10.1016/J.JACL.2017.04.114

19. Wilson DP, Jacobson TA, Jones PH, Koschinsky ML, McNeal CJ, Nordestgaard BG, Orringer CE. Use of Lipoprotein(a) in clinical practice: A biomarker whose time has come. A scientific statement from the National Lipid Association. J Clin Lipidol. 2019;13(3):374–392. doi:10.1016/J.JACL.2019.04.010

20. Enkhmaa B, Anuurad E, Berglund L. Lipoprotein (a): impact by ethnicity and environmental and medical conditions. J Lipid Res. 2016;57(7):1111–1125. doi:10.1194/jlr.R051904

21. Reyes-Soffer G. The impact of race and ethnicity on lipoprotein(a) levels and cardiovascular risk. Curr Opin Lipidol. 2021;32(3). https://journals.lww.com/co-lipidology/Fulltext/2021/06000/The_impact_of_race_and_ethnicity_on_lipoprotein_a_.3.a spx

22. Tsimikas S, Marcovina SM. Ancestry, Lipoprotein(a), and Cardiovascular Risk Thresholds: JACC Review Topic of the Week. J Am Coll Cardiol. 2022;80(9):934–946. doi:10.1016/J.JACC.2022.06.019

23. Saleheen D, Haycock PC, Zhao W, Rasheed A, Taleb A, Imran A, Abbas S, Majeed F, Akhtar S, Qamar N, Zaman KS, Yaqoob Z, Saghir T, Rizvi SNH, Memon A, Mallick NH, Ishaq M, Rasheed SZ, Memon F ur R, Mahmood K, Ahmed N, Frossard P, Tsimikas S, Witztum JL, Marcovina S, Sandhu M, Rader DJ, Danesh J. Apolipoprotein(a) isoform size, lipoprotein(a) concentration, and coronary artery disease: a mendelian randomisation analysis. Lancet Diabetes Endocrinol. 2017;5(7):524–533. doi:10.1016/S2213-8587(17)30088-8

24. Hanif S, Akhtar B, Afzal MN. Serum lipoprotein (A) levels in acute coronary syndrome; comparison of younger and elderly patients with healthy controls. Pak J Med Sci. 2019;35(6):1718–1723. doi:10.12669/pjms.35.6.377

25. Habib SS, Aslam M. Lipids and lipoprotein(a) concentrations in Pakistani patients with type 2 diabetes mellitus. Diabetes Obes Metab. 2004;6(5):338–343. doi:10.1111/j.1462-8902.2004.00352.x

26. Paré G, Çaku A, McQueen M, Anand SS, Enas E, Clarke R, Boffa MB, Koschinsky M, Wang X, Yusuf S. Lipoprotein(a) Levels and the Risk of Myocardial Infarction Among 7 Ethnic Groups. Circulation. 2019;139(12):1472–1482. doi:10.1161/CIRCULATIONAHA.118.034311

27. Sadiq F, Shafi S, Sikonja J, Khan M, Ain Q, Khan MI, Rehman H, Mlinaric M, Gidding SS, Groselj U, Alam J, Ali M, Anwer J, Awan WA, Bham SQ, Fatima N, Gul F, Hameed SS, Haroon M, Hasan M, Jadoon A, Jamil S, Khan AA, Khan SA, Kidwai SS, Munir A, Bin Nazir MT, Khan Niazi GZ, Qabulio SN, Rana MA, Rehman A ur, Safdar S, Shah S, Rehman Ahmed Sheikh TU, Yousuf A, Zehra K, Zehra T. Mapping of familial hypercholesterolemia and dyslipidemias basic management infrastructure in Pakistan: a cross-sectional study. The Lancet Regional Health - Southeast Asia. 2023;0(0):100163. doi:10.1016/j.lansea.2023.100163

28. Kelsey MD, Mulder H, Chiswell K, Lampron ZM, Nilles E, Kulinski JP, Joshi PH, Jones WS, Chamberlain AM, Leucker TM, Hwang W, Milks MW, Paranjape A, Obeid JS, Linton MF, Kent ST, Peterson ED, O’Brien EC, Pagidipati NJ. Contemporary patterns of lipoprotein(a) testing and associated clinical care and outcomes. Am J Prev Cardiol. 2023;14:100478. doi:10.1016/j.ajpc.2023.100478

29. Laufs U, Schorr J, Klebs S. Characteristics of patients with a lipoprotein(a) assessment – a health insurance claims database analysis. Eur Heart J. 2021;42(Supplement_1):ehab724.2519. doi:10.1093/eurheartj/ehab724.2519

30. Stürzebecher PE, Schorr JJ, Klebs SHG, Laufs U. Trends and consequences of lipoprotein(a) testing: Cross-sectional and longitudinal health insurance claims database analyses. Atherosclerosis. 2023;367:24–33. doi:10.1016/j.atherosclerosis.2023.01.014

31. Zafrir B, Aker A, Saliba W. Lipoprotein(a) testing in clinical practice: real-life data from a large healthcare provider . Eur J Prev Cardiol. 2022;29(14):e331–e333. doi:10.1093/eurjpc/zwac124

32. Nissen SE, Wolski K, Cho L, Nicholls SJ, Kastelein J, Leitersdorf E, Landmesser U, Blaha M, Lincoff AM, Morishita R, Tsimikas S, Liu J, Manning B, Kozlovski P, Lesogor A, Thuren T, Shibasaki T, Matei F, Silveira FS, Meunch A, Bada A, Vijan V, Bruun NE, Nordestgaard BG. Lipoprotein(a) levels in a global population with established atherosclerotic cardiovascular disease. Open Heart. 2022;9(2). doi:10.1136/openhrt-2022-002060

33. Dahlen GH, Guyton JR, Attar M, Farmer JA, Kautz JA, Gotto AM. Association of levels of lipoprotein Lp(a), plasma lipids, and other lipoproteins with coronary artery disease documented by angiography. Circulation. 1986;74(4):758–765. doi:10.1161/01.CIR.74.4.758

34. Varvel S, McConnell JP, Tsimikas S. Prevalence of Elevated Lp(a) Mass Levels and Patient Thresholds in 532 359 Patients in the United States. Arterioscler Thromb Vasc Biol. 2016;36(11):2239–2245. doi:10.1161/ATVBAHA.116.308011

35. Nicholls SJ, Tang WHW, Scoffone H, Brennan DM, Hartiala J, Allayee H, Hazen SL. Lipoprotein(a) levels and long-term cardiovascular risk in the contemporary era of statin therapy. J Lipid Res. 2010;51(10):3055–3061. doi:10.1194/jlr.M008961

36. Werba JoséP, Safa O, Gianfranceschi G, Michelagnoli S, Sirtori CR, Franceschini G. Plasma triglycerides and lipoprotein(a): inverse relationship in a hyperlipidemic Italian population. Atherosclerosis. 1993;101(2):203–211. doi:10.1016/0021-9150(93)90117-D

37. Bartens W, Rader DJ, Talley G, Brewer Jr. HB. Decreased plasma levels of lipoprotein(a) in patients with hypertriglyceridemia. Atherosclerosis. 1994;108(2):149–157. doi:10.1016/0021-9150(94)90109-0

38. Konerman M, Kulkarni K, Toth PP, Jones SR. Evidence of dependence of lipoprotein(a) on triglyceride and high-density lipoprotein metabolism. J Clin Lipidol. 2012;6(1):27–32. doi:10.1016/j.jacl.2011.08.004

39. McConathy WJ, Trieu VN, Koren E, Wang CS, Corder CC. Triglyceride-rich lipoprotein interactions with Lp(a). Chem Phys Lipids. 1994;67-68:105–113. doi:10.1016/0009-3084(94)90129-5

